# A High Through-put Assay for Circulating Antibodies Directed against the S Protein of Severe Acute Respiratory Syndrome Coronavirus 2 (SARS-CoV-2)

**DOI:** 10.1101/2020.04.14.20059501

**Authors:** Svenja Weiss, Jéromine Klingler, Catarina Hioe, Fatima Amanat, Ian Baine, Erna Milunka Kojic, Jonathan Stoever, Sean Liu, Denise Jurczyszak, Maria Bermudez-Gonzalez, Viviana Simon, Florian Krammer, Susan Zolla-Pazner

## Abstract

**Background:** More than one million infections with the severe acute respiratory syndrome corona virus 2 (SARS-CoV-2) have been confirmed. While PCR-based assays are used for diagnosis, high through-put serologic methods are needed to detect antibodies for seroserveillance and for identification of seroconversion, potential plasma donors, and the nature of the immune response to this pathogen.

**Methods:** A Luminex binding assay was used to assess the presence of antibodies in human sera from COVID-19-infected and -uninfected individuals specific for two recombinant proteins of SARS-CoV-2.

**Findings:** Fluorochrome-labeled beads were coated with a recombinant soluble stabilized trimeric SARS-CoV-2 S protein ectodomain or its central portion, the receptor binding domain (RBD). Coated beads were incubated with sera, followed by incubation with biotinylated anti-human total Ig antibodies and phycoerythrin (PE)-labeled streptavidin. Readout using a Luminex analyzer clearly differentiated between sera of the infected and uninfected subjects, delineating a wide range of serum antibody levels in infected subjects.

**Interpretation:** Antibody assays of sera can identify individuals who are infected with SARS-CoV-2 and have seroconverted, as well as subjects who have been infected and recovered. The use of the Luminex binding Ab assay has the advantage that it can be run in approximately 2.5 hours, uses very little antigen, and permits a high through-put of samples/day.

**Funding:** NIAID contracts and grants, Department of Veterans Affairs’ grants, the Microbiology Laboratory Clinical Services, Translational Science Hub, and Personalized Virology Initiative, and Department of Medicine of Mount Sinai Health System and Icahn School of Medicine at Mount Sinai.

**RESEARCH IN CONTEXT:** *Evidence before this study:* The outbreak of infections with SARS-CoV-2 began in late 2019. Specimens from nasopharyngeal swabs are being used in PCR-based assays to test for the presence of the virus. Until the first week in April, 2020 there were no licensed tests for the presence of serum antibodies against proteins of the virus. The first approved tests are now becoming available, but none use a format that can be scaled up for mass screening which is now needed for implementing various public health measures. As per a recent Pubmed search, less than 10 studies using serologic assays have been published and none are high through-put.

*Added value of this study:* High through-put antibody tests are needed in order to identify seroconversion, to perform serosurveys, identify potential donors for plasma therapy, assess the prevalence of infection in populations, identify healthcare workers who may be immune to SARS-CoV-2, and to study the nature of the immune response to this pathogen. The method described for detecting antibodies in SARS-CoV-2-infected patients can be applied in hospital and reference labs, allowing the assessment of present and past infection in a much higher number of donors per unit of time than assays described heretofore.

*Implications of all the available evidence:* This study shows that a test in which magnetic beads are coated with soluble forms of the spike protein of SARS-CoV-2 can be used to test for the presence of antibodies targeting this pathogen. The platform allows for the efficient testing of multiple specimens simultaneously using as little as 5 nanograms of antigen per test. This test affords the possibility of large scale, economical and efficient antibody testing.

## INTRODUCTION

The reverse transcription polymerase chain reaction (RT-PCR) test is currently being used for the qualitative detection of SARS-CoV-2 nucleic acids in specimens from the upper and lower respiratory tract.^1,2^ Molecular testing is well established and has been used in clinical laboratories throughout the world for two decades. In contrast, testing of serum and other bodily fluids for antibodies (Abs) to infectious diseases such as syphilis, typhoid and diphtheria have been used for over a century.^3,4^ Antibody (Ab) assays are most useful for identifying individuals who have been infected with a particular pathogen and seroconverted. As such, they can be particularly valuable, for example, for identification of subjects who have had asymptomatic viral infections and those who have recovered and would no longer be positive in tests for viral nucleic acids. They would also be particularly useful for serosurveillance, to identify donors for COVID-19 plasma therapy, and to identify individuals who are potentially immune to reinfection. Antibody assays thus fill an essential gap both during and after the current SARS-CoV-2 pandemic. In fact, in one study, depending on the time of testing post-infection, the combined use of RT-PCR and Ab positivity provided an advantage over either test alone.^5^ We and others^5-7^ have described tests for assessing the presence of Abs to SARS-CoV-2 in serum and plasma using the enzyme-linked immunosorbent assay (ELISA) platform with a recombinant form of the S protein of the virus and/or the central portion of this molecule identified as the receptor binding domain (RBD), consisting of amino acids 319-541.^7-9^ We report here a modification of the ELISA assay in which beads labeled with a particular fluorochrome signature are coated with the soluble recombinant S protein or RBD, incubated with serum, biotinylated anti-human total Ig Abs, and phycoerythrin (PE)-labeled streptavidin. The readout is performed with a laser-based instrument. This is a high through-put assay that offers the advantages of being able to prepare the antigen-coated beads for thousands of tests in a single day and using at least 20-fold less antigen than is required for ELISA. In the setting of hospitals and regional reference labs, results on >5,000 specimens per day can be generated.

## METHODS

### Recombinant proteins

The recombinant S and RBD proteins were produced as previously described^7^ in Expi293F cells (ThermoFisher) by transfections of purified DNA using an ExpiFectamine Transfection Kit (ThermoFisher). The soluble version of the spike protein included the S protein ectodomain (amino acids 1-1213), a C-terminal thrombin cleavage site, a T4 foldon trimerization domain and a hexahistidine tag. The protein sequence was modified to remove the polybasic cleavage site (RRAR to A) and two stabilizing mutations (K986P and V987P, wild type numbering). The RBD (amino acids 319-541) also contained a hexahistidine tag. Supernatants from transfected cells were harvested on day three post-transfection by centrifugation of the culture at 4000 g for 20 minutes. Supernatant was then incubated with 6 mL Ni-NTA agarose (Qiagen) for one to two hours at room temperature. Next, gravity flow columns were used to collect the Ni-NTA agarose and the protein was eluted. Each protein was concentrated in Amicon centrifugal units (EMD Millipore) and re-suspended in phosphate buffered saline (PBS).

### Human samples

Banked serum samples were obtained from study participants enrolled in two IRB-approved longitudinal observational protocols (Icahn School of Medicine at Mount Sinai; PI: Dr. V. Simon: IRB-16-00772 and IRB-16-00791). Samples from four participants with documented SARS-CoV-2 infection were available for P#1 (three time points), P#2 (two time points), and P#3 and P#4 (one time point each). Sera collected from healthy donors in the months of October and November in 2019, prior to the spread of SARS-Cov-2 in the USA, served as negative controls (N#1, N#2, N#3). All participants provided written consent at study enrollment and agreed to sample banking and future research use of their banked biospecimen. Four additional de-identified serum specimens (P#5-8) were provided by the Clinical Pathology Laboratory at the Icahn School of Medicine at Mount Sinai. **Table 1** provides an overview of the serum samples used in this report.

**Table 1.** Characteristics of patients whose sera were tested.

| Pt. # | SARS-Cov-2<br>PCR Result | Symptoms<br>suggestive<br>of COVID-19 | Days post<br>symptom onset<br>and serum<br>collection | COVID19<br>severity |
| --- | --- | --- | --- | --- |
| P#1a | positive | Yes | 8 | severe |
| P#1b | positive | Yes | 11 | severe |
| P#1c | positive | Yes | 15 | severe |
| P#2a | positive | Yes | 7 | severe |
| P#2b | positive | Yes | 10 | severe |
| P#3 | positive | Yes | 22 | mild |
| P#4 | inconclusive | Yes | 21 | mild |
| P#5* | positive | unknown | unknown | unknown |
| P#6* | positive | unknown | unknown | unknown |
| P#7* | positive | unknown | unknown | unknown |
| P#8* | positive | unknown | unknown | unknown |
| N#1 | n/a; collected Oct. 2019 | n/a | n/a | n/a |
| N#2 | n/a; collected Nov. 2019 | n/a | n/a | n/a |
| N#3 | n/a; collected Oct. 2019 | n/a | n/a | n/a |

\*Patients specimens P#5-P#8) were de-identified
n/a = not applicable

### Luminex binding Ab assay

The SARS-CoV-2 antigens included a soluble recombinant trimerized form of the S protein and a recombinant RBD protein produced in Expi293F mammalian cells as described (mSpike, mRBD).^7^ The antigens were covalently coupled individually to carboxylated xMAP beads at 4.0 μg protein/million beads using a two-step carbodiimide reaction with the xMAP Ab Coupling (AbC) Kit according to the manufacturers’ instructions (Luminex, Austin, TX). The coupled beads are pelleted, resuspended in storage buffer at 5 × 10^6^/mL (PBS, 0.1% bovine serum albumin (BSA), 0.02% Tween-20, and 0.05% sodium azide, pH 7.4), and stored at −80° C. Three to five million beads were prepared in a single 1.5 mL conical tube.

The beads needed for a single run (2,500 beads/well x number of wells) were pelleted and resuspended in assay buffer (PBS, 0.1% BSA, 0.02% Tween-20) to a volume that delivered 2,500 beads to each well in an aliquot of 50 μL/well. Serum was diluted in assay buffer to twice the desired dilution, added as 50 μL/well to the wells containing the beads and incubated at room temperature for 1 hr on a plate shaker at 600 rpm. After two washes in assay buffer, 100 μL/well of biotinylated-anti-human total Ig (Abcam, catalog #ab97158) at 2 μg/ml was added and incubated for 30 min at room temperature on a plate shaker. After two washes, 100 μL/well of Streptavidin*-*Phycoerythrin (PE) at 1 μg/ml was added (BioLegend Catalog #405204) followed by a 30 min incubation at room temperature on a plate shaker. After two additional washes, 100 μL of assay buffer/well was added and put on a shaker to resuspend the beads. The plate was read with a Luminex Flexmap 3D instrument. Samples were tested in duplicate and the results were recorded as mean fluorescent intensity (MFI). Serum was used for the experiments herein. In previous work using a similar format, both serum and plasma have been used (unpublished data). Sera from uninfected individuals, drawn before the start of the COVID-19 epidemic served as negative controls. Figures were generated using GraphPad Prism 7.02.

## RESULTS

Eleven serum specimens were tested from eight different patients with confirmed SARS-CoV-2 infection. Specimens from four uninfected individuals were used which had been banked as part of an ongoing longitudinal study prior to the COVID-19 outbreak. The assays were performed in a BSL 2 lab since they do not involve live virus; normal blood precautions employed when handling all human specimens were used for these studies. Serum specimens were heat inactivated (30 min at 60 C) prior to use in the assay. The specimens were screened for the presence of Abs reactive with soluble recombinant trimeric mammalian cell expressed S protein (mSpike) or mRBD (**Figure 1**) using a secondary Ab that detects total Ig. All patient specimens reacted strongly with SARS-CoV-2 antigens (**Figures 1** and **2**), although reactivity with the mSpike was generally stronger than with mRBD (**Figure 1**). This is consistent with the findings of Amanat et al.^7^ although different sera were used in their experiments and ours. The three sera from the uninfected patients did not react with either antigen (**Figures 1** and **2**), and there was no reactivity in the presence of PBS but no serum (data not shown).

**Figure 1.**
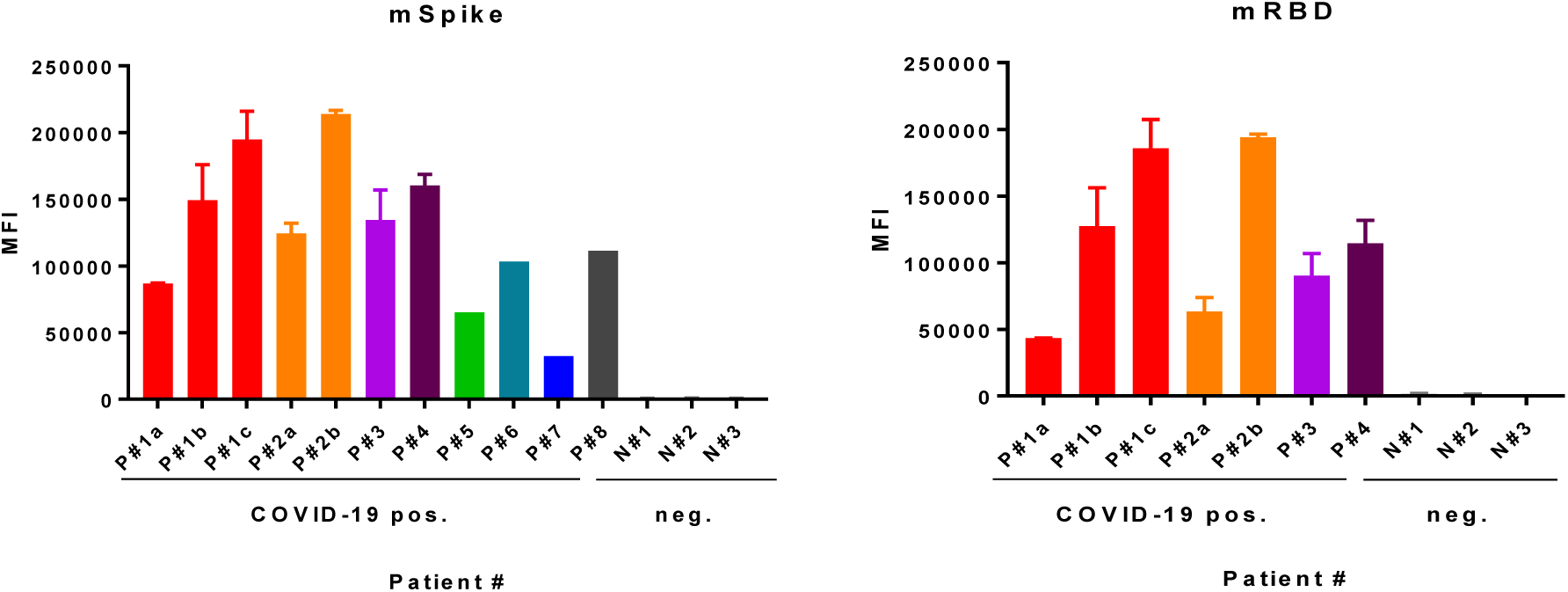
Screening for the presence of SARS-CoV-2 antibodies in specimens from COVID-19-infected and uninfected (neg) humans. Assays were run using the S protein produced in mammalian cells (mSpike, left) with sera from eight infected patients and with sera from four of these patients with mRBD (right). Results are shown using sera tested at a dilution of 1:200. For specimens from four patients (P#1, 2, 3 and 4) run against both antigens, the data shown are the mean + S. D. of two to five experiments. For four patients (P#5-8), the specimens were run only against the mSpike in a single experiment.

**Figure 2.**
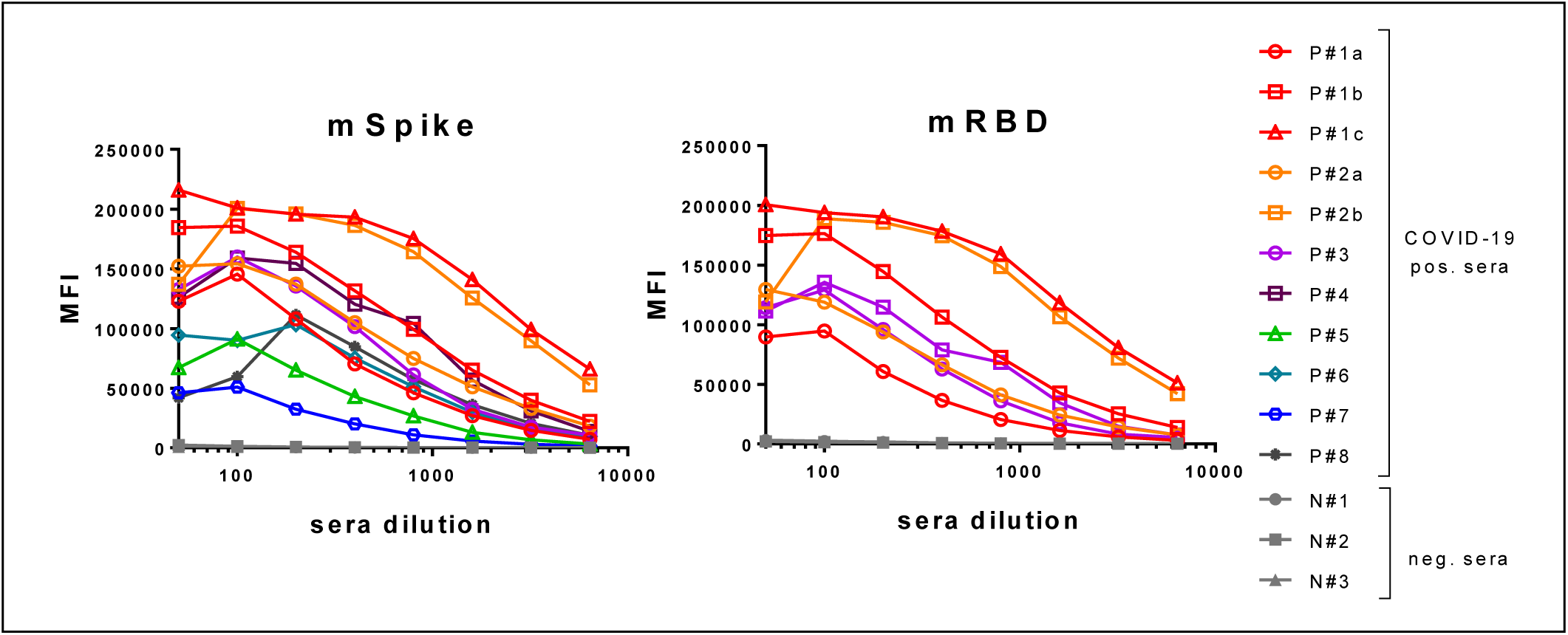
Titration of COVID-19-positive and negative sera. Specimens were diluted at 2-fold dilutions from 1:50 - 1:6,400 and tested using the Luminex assay described herein. Titration curves are shown for sera from 11 specimens from eight infected patients using the mSpike as antigen (left) and for sera from three patients tested vs. mRBD (right).

Notably, this study was not designed as a Diagnostic Accuracy Study; at least 40 or more specimens would need to be included to establish a significant cut-off value; however it is striking that all 11 PCR positive specimens were Ab positive with MFI values >25,000 and all 3 PCR negative specimens were Ab negative with MFI values <2,000.

To determine the dynamic range of the assay and determine the titer of Abs present in COVID-19-positive sera, titrations were performed with all 11 specimens using the mSpike antigen, and with seven of these specimens using the mRBD antigen; the three specimens from the uninfected subjects were titrated against both antigens. The titration curves are shown in **Figure 2**. A prozone effect (the phenomenon in which a value below the peak is observed at low dilutions of sera) is suggested by the curves generated by six of the 11 sera tested against the mSpike. The prozone phenomenon was less apparent when using the mRBD antigen. Generally the maximum MFI values were achieved with serum dilutions of 1:100 or 1:200. Levels of Abs varied greatly between the specimens tested. For example, MFI values at a serum dilution of 1:200 from infected patients ranged from 25,000 to >200,000 (**Figure 1**). Of interest, titration curves show that the specimens with the highest Ab levels (P#1c and P#2b) were from two different patients (P#1 and P#2), and these two subject were the two patients with severe disease at the time of the blood draw. (**Table I**). Further study is needed to ascertain how and whether disease severity affects the level and type of Abs induced. However, it is notable that the level of Abs increased over a period of a few days in each of the two patients from whom we had longitudinal specimens (P#1 and P#2, **Table I**). The factors that affect the variation in Ab responses (severity of disease, length of infection, gender, genetics, etc) are yet to be determined and will require a large panel of specimens from patients with adequate clinical and demographic data.

## DISCUSSION

A rapid through-put test for SARS-CoV-2 Abs, such as the Luminex bead binding Ab assay described here, can serve as an essential tool for identifying infected individuals, complementing the PCR-based methods measuring the presence and level of SARS-CoV-2. The Ab assay will be a particularly useful assay for identifying individuals who have had asymptomatic infections, including children.^10-12^ Given the relatively high prevalence of asymptomatic individuals who have or have had SARS-CoV-2 infection, Ab assays thus fill an essential gap both during and after the current SARS-CoV-2 pandemic since the Luminex Ab assay can be performed in approximately 2.5 hr (**Figure 3**) with extremely small amounts of antigen per test, in a platform that can process thousands of samples per day.

**Figure 3.**
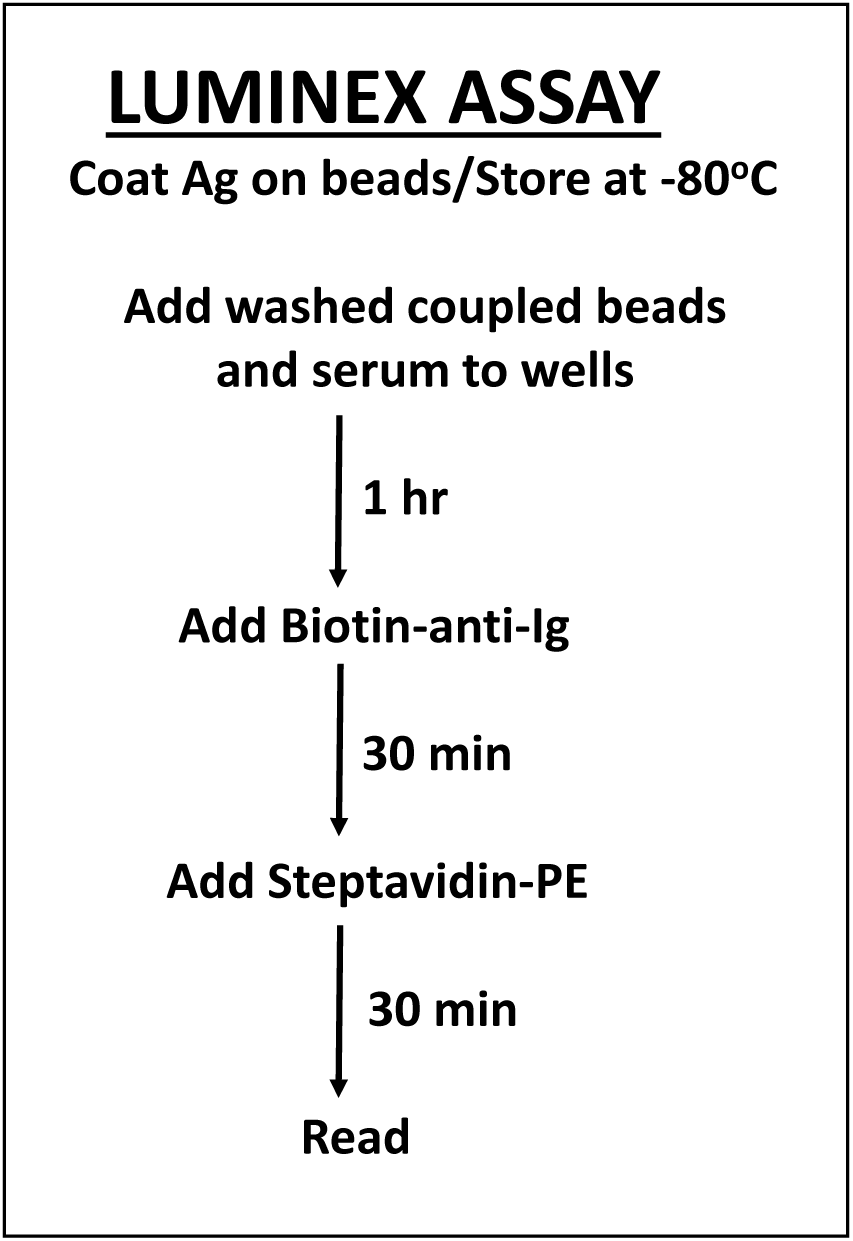
Steps and time required for detection of antibodies to the mSpike or mRBD of SARS-CoV-2 antigens using the Luminex antibody binding assay described herein. Washing steps, delineated in Methods, add approximately 30 minutes, resulting in a test requiring approximately 2.5 hr.

The Ab assay described here uses antigen-coated magnetic beads that can be prepared in bulk and used for at least four weeks, simplifying the work load and logistics in the laboratory. It is worth noting that, using various HIV proteins and peptides, we have used antigen-coated xMAP beads stored under sterile conditions as described in Methods, for up to 4 weeks.^13^ The length of time that xMAP beads coated with the mSpike or mRBD proteins can be stored and used successfully has not yet been determined, but barring degradation of the proteins at −80° C under sterile conditions, it is anticipated that they would be stable and useful for 4 weeks or longer. This means that the antigen-coated beads could be prepared and stored in batches providing reagents for several thousands of tests.

A concentration of 4 μg/10^6^ beads was used for bead coating in the experiments described above since that concentration had been used previously with various proteins; a concentration of 2 μg/10^6^ beads gave comparable values but <2 μg/10^6^ beads gave lower MFI values in titration curves with sera from infected individuals (data not shown). Given that the S and RBD recombinant proteins are currently in limited supply, the minimum concentration for bead-coating is advantageous, with coating at 4 and 2 ug/million beads resulting in the use of 0.01 and 0.005 μg/test, respectively—a level considerably less than that required for most ELISA assays. Further modification is underway to determine if other bodily fluids, such as urine and/or saliva, can replace serum in this assay. Both urine and saliva Ab tests are used for diagnostics in other diseases such as HIV and TB.^14,15^

This assay can be scaled up for use in hospital and reference laboratories. A high through-put assay such as this can be used for a multitude of purposes including serosurveillance of communities, screening of health care workers who may have developed immunity to SARS-CoV-2, identification of donors for COVID-19 plasma therapy, and selection of individuals who are potentially immune to reinfection.

## Data Availability

All data referred to in the manuscript are available to the research and clinical community.

## ACKNOWLEDGMENTS

We are extremely grateful to the COVID-19 patients for their contribution to research and wish for them a full recovery.

